# Full-Field Stimulus Test for Visual Function Assessment in Ultra-Low Vision with Retinitis Pigmentosa

**DOI:** 10.64898/2026.04.26.26350738

**Authors:** Lizhu Yang, Yusaku Katada, Kaoru Fujinami, Shiori Yamamoto, Konomi Fukuda, Ari Shinojima, Yohei Tomita, Norimitsu Ban, Hiromitsu Kunimi, Hajime Shinoda, Kazuno Negishi, Toshihide Kurihara

## Abstract

**Purpose:** Assessing visual function in patients with ultralow vision (ULV), particularly those with retinitis pigmentosa (RP), remains a significant challenge in therapeutic development. Full-field stimulus test (FST) provides a quantitative measure of retinal light sensitivity and may serve as a valuable clinical endpoint. We investigated FST in ULV RP by examining its associations with functional measures and daily activity–based tasks.

**Design:** Observational, cross-sectional study.

**Participants:** Patients with RP and visual acuity in the worse-seeing eye below counting fingers (CF) were enrolled.

**Methods:** After dilation and 45-minute dark adaptation, FST was performed monocularly with brief full-field white-light flashes across three visits. Visual acuity was classified into four groups: no light perception (NLP), light perception (LP), hand motion (HM), and CF or better. We assessed functional vision using two tabletop object-recognition and exploration tasks, two mobility tasks, and three vision-related questionnaires. FST thresholds were compared across visual acuity groups, and associations with functional outcomes were analyzed.

**Main Outcome Measures:** FST thresholds and their associations with functional vision outcomes.

**Results:** Thirty-five patients (70 eyes; median age, 62 years, range 39–84) were included. Median FST thresholds (log cd⋅s/m²) by visual acuity group were as follows: NLP, 1.13 (-0.63–2.54); LP, -0.27 (-2.70–2.91); HM, -1.13 (-6.24–0.51); CF or better, -2.82 (-5.67– -1.73) (p < 0.001). Measurable FST thresholds were obtained in 9 of 14 NLP eyes (64.3%). FST thresholds showed significant correlations with tabletop performance (r = –0.70 to – 0.46) and mobility performance (r = –0.65), whereas no significant association was observed with questionnaire scores. Test–retest variability across three visits showed no systematic bias, with a coefficient of repeatability of ±0.66 to ±0.82 log cd·s/m². ROC analyses identified FST cutoffs of –1.75 to –0.87 log cd·s/m² at which patients first achieved nonzero functional task performance.

**Conclusions:** FST quantifies residual visual function in ULV RP and correlates strongly with performance-based measures of functional vision in daily life. These findings support FST as a clinically meaningful endpoint for therapeutic trials in advanced RP and other severe visual impairments and highlight the value of anchoring FST thresholds to functional task performance.

## Introduction

Retinitis pigmentosa (RP) is a group of inherited retinal degenerative disorders characterized by the progressive loss of rod and cone photoreceptors. The prevalence of RP is estimated at approximately 1 in 4000 individuals, making RP one of the leading causes of severe visual impairment worldwide. In typical RP, rod photoreceptor degeneration occurs before cone degeneration, resulting in early symptoms such as night blindness and progressive constriction of the visual field, followed by central vision loss and, in advanced cases, complete blindness^1,2^.

In recent years, novel therapeutic strategies—including gene-based and stem cell-derived therapies—have emerged as potential treatments for RP^3–7^. These approaches target different levels of the visual pathway, highlighting the critical need for effective clinical trial endpoints that can quantitatively and accurately assess visual function. Conventional visual acuity (VA) testing remains the most commonly used clinical measure; nevertheless, it is insufficient for patients with ultra-low vision (ULV), defined as those categorized with off-chart VA levels, such as counting fingers (CF), hand motion (HM), light perception (LP), or no light perception (NLP). These patients are the primary candidates for these new treatments. Traditional VA tests fail to quantify residual visual function in these patients and cannot detect subtle changes following therapeutic intervention^8^. To overcome this limitation, alternative endpoints such as the full-field stimulus test (FST) and multi-luminance mobility tests have been introduced in clinical trials^7^. The multi-luminance mobility test more directly reflects real-life visual function; however, it is more costly and difficult to set up a mobility course in most clinical environments. FST has a greater dynamic range, is easier to implement in clinical settings, and requires minimal space. FST utilizes full-field light stimuli of different wavelengths and intensities under dark- and light-adapted conditions to estimate the threshold at which a patient perceives light. Unlike visual acuity tests, FST does not require spatial resolution or fixation, making it particularly suitable for patients with ULV. Furthermore, FST can theoretically differentiate rod- and cone-mediated responses, allowing insight into the functional status of photoreceptor subtypes^9–11^. Notably, FST can detect residual light sensitivity even in patients categorized as NLP or LP, whereas conventional VA testing provides no quantifiable information^11,12^. This ability is particularly critical, as these patients often represent the primary target population for novel therapies such as optogenetics and stem cell–based interventions. FST is not only a research tool but has been increasingly recognized as a candidate endpoint for interventional trials in ultra-low vision. Its scalability, relatively short testing time, and minimal space requirements make it feasible for multicenter studies^10,11^. In Japan, based on findings from the US Phase 3 Luxturna trial demonstrating a correlation between FST and MLMT^7^, FST was adopted in 2019 as a primary outcome measure in a Phase 3 trial of RPE65 gene therapy. This study design, including the use of FST as a primary endpoint, was approved by the Pharmaceuticals and Medical Devices Agency (PMDA), marking the first such application worldwide^4^. Additionally, recent international guidelines from the International Society for Clinical Electrophysiology of Vision (ISCEV) and the Imaging and Perimetry Society (IPS) have provided the guideline, facilitating their acceptance by regulators as a reliable and reproducible outcome measure in IRD trials^9^.

Most therapeutic trials have enrolled patients with relatively preserved visual acuity and retinal structure; however, the use of FST in ULV patients with advanced RP remains insufficiently characterized. In particular, the relationship between FST thresholds and functional vision in daily life remains underinvestigated. By linking FST to performance-based measures—such as mobility and tabletop object-recognition tasks—its outcomes can be anchored to everyday activities (locating tableware on a surface or walking toward a doorway), thereby providing clinical relevance beyond psychophysical sensitivity alone^7,13^.

In this study, we aimed to evaluate the utility of FST in patients with ULV, focusing on its correlation with other functional and structural measures and its relevance to daily life activities. The findings of this study may help determine whether FST can serve as a meaningful efficacy endpoint for assessing visual function changes in clinical trials targeting patients with ULV.

## Methods

### Patients

This single-center, prospective observational study was approved by the Institutional Review Board of Keio University School of Medicine (approval reference: 20211118) and conducted in accordance with the Declaration of Helsinki. Written informed consent was obtained from all participants before data collection. Patients diagnosed with RP who visited Keio University Hospital (Tokyo, Japan) between November 2022 and November 2024 were recruited. Inclusion criteria required the VA of the worse eye to be equal to or worse than CF (≥ LogMAR 2.0 [equivalent to 0.01 decimal]). Patients with other ocular diseases that could affect visual function, such as severe cataract, corneal disease, or optic nerve disorders, were excluded.

All participants underwent a series of examinations, including best-corrected VA, intraocular pressure measurement, slit-lamp and fundus examination, spectral-domain optical coherence tomography (Heidelberg Engineering, Spectralis), FST, mobility tests, table tests, and visual function questionnaires. We performed visual function tests and FST at three separate visits, each spaced at intervals of at least 1 week to ensure reproducibility.

### VA Measurements

We measured best-corrected VA using the Early Treatment Diabetic Retinopathy Study letter chart (Precision Vision, Bloomington, IL, USA). When a patient was unable to correctly identify three or more letters on the first line of the chart at 50 cm, we assessed off-chart VA using standardized criteria: CF, HM, LP, and NLP. CF was recorded if the patient could correctly count the number of the examiner’s fingers presented at a distance of 2 feet. If the patient could not count fingers, we assessed HM by moving a hand 2 feet in front of the patient and recorded it as HM if hand movement was detected. If HM was not detected, we tested LP by directing light from a monocular indirect ophthalmoscope into the patient’s pupil in a darkened room. We recorded VA as LP when the patient perceived light and as NLP when the patient did not^14^.

### FST

FST was performed using the Espion Ganzfeld Profile System (Diagnosys LLC, Boston, MA, USA). Patients were pharmacologically dilated and dark-adapted for 45 minutes prior to testing. Stimuli consisted of 4-ms white light flashes. Testing was conducted monocularly, with the fellow eye occluded. Before the actual FST measurements, patients completed a practice session to minimize learning effects. During testing, patients responded using a textured two-button box (“yes” or “no”) to indicate whether they perceived the stimulus. We defined the FST threshold as the midpoint of the frequency-of-seeing curve, derived from a two-parameter Weibull function that accounted for false-positive and false-negative responses^4^.

We repeated FST assessments 4 times per eye, with up to 6 attempts if all previous assessments yielded a quality score of 0. We considered a measurement reliable when at least one assessment had a quality score of ≥ 1, as calculated automatically by the system. We determined the final FST threshold for each eye as the mean of all reliable assessments.

### Table Test

We performed two table tests to assess visual function in relation to activities of daily living (ADL). We adapted Table Test A from a previously described protocol used in subretinal retinal prosthesis studies^15^. We placed white items—comprising one set of four types of tableware and a second set of four distinct geometric objects—on a homogeneously illuminated black tablecloth. We instructed patients to report the number, locations, and types of items. For the tableware task, each trial was scored on number (2 points if correct), location (1 point per item), and type (1 point per item); the final score was the mean across six trials. For the figure task, each trial scored 1 (correct identification) or 0; the final score was the proportion correct across six trials (range, 0–1).

We adapted Table Test B from the GS030-DP optogenetics clinical trial^6^. The first part of this test, termed the visual perception test, assessed eye–hand coordination. We selected one object from different types with varying contrast levels, placed it on a white tablecloth, and asked the patient to locate and touch it. The second part, termed visual exploration and relative position, involved placing three cups with different contrast levels in six possible positions on the table. We asked patients to report the number of objects perceived and their relative positions. To reflect the visual function of daily life, we performed all table tests binocularly. For each test, we recorded scores and completion time as outcome measures. The visual perception test scored each trial on recognition, position, and touch (range, 0–3 per trial); the visual exploration test scored number and position (range, 0–3 or 0–4 per trial depending on the number of objects). Final scores were the means across six trials. Further methodological details are provided in the Supplemental Information.

### Mobility Tests

We adapted two mobility tasks from prior studies of suprachoroidal retinal prostheses^16,17^. We conducted the tests in an illuminance-controlled room with white walls and a black floor, which included a *Door Task* and a Line Task. In the Door Task, we positioned a simulated door made of black cloth on a white wall across the room; we instructed patients to locate the door, walk toward it, and touch it. In the Line Task, we drew a white line on a black floor and instructed patients to follow the line to its end.

We performed all mobility tasks binocularly under six ambient illuminance levels (400, 250, 100, 50, 10, and 1 lx). A trial was considered successful if the participant reached and touched the door (Door Task) or stopped within 30 cm of the line’s distal end with at least part of one foot on the line (Line Task). Participants had to pass all six trials at a given illuminance level to proceed to the next lower level. For each task, we converted the minimum illuminance passed into an ordinal score ranging from 0 (unable to pass at 400 lx) to 6 (passed at 1 lx). We recorded both the score and task completion time as outcome measures. Further methodological details are provided in the Supplemental Information.

### Vision-Related Questionnaire

To evaluate vision-related quality of life (QoL), we used three interviewer-administered questionnaires: the 25-item National Eye Institute Visual Function Questionnaire (NEI VFQ-25)^18^, the Impact of Vision Impairment Questionnaire (IVI)^19^, and the Impact of Vision Impairment–Very Low Vision (IVI-VLV)^20^. We administered all questionnaires in Japanese. Given that most participants were unable to read the items, trained examiners read each question aloud and recorded patient responses. We calculated scores (and subscale scores, where applicable) according to each instrument’s scoring guidelines.

### Definition of terms

In this study, the term *functional vision* refers specifically to performance on structured, activity-based tasks (Mobility and Table Test) that approximate the use of residual vision in real-world contexts. The term “*daily activities”* refers to basic tasks reflected by these measures, such as object recognition and navigation, which are directly relevant to patient independence and safety. Throughout the manuscript, *daily activity–based assessments* specifically denote the Table and Mobility Tests, which we designed to model ADL-relevant functions under standardized conditions.

### Statistical Analysis

We performed all analyses using SPSS version 30 (IBM Corp., Armonk, NY, USA). We summarized continuous variables as mean ± SD or median (range), depending on the distribution of the data. We assessed normality with the Shapiro–Wilk test, and homogeneity of variances with Levene’s test. We examined group differences in FST thresholds across visual acuity categories using one-way ANOVA when parametric assumptions were met, or the Kruskal–Wallis test otherwise. When overall group differences were significant, we performed post-hoc comparisons using Tukey’s honestly significant difference test for ANOVA and Dunn’s test with Bonferroni adjustment for Kruskal–Wallis analyses. We assessed associations between FST thresholds and results from other examinations using Pearson’s correlation for approximately normally distributed variable pairs and Spearman’s rank correlation otherwise. All statistical tests were two-sided, and we considered *p*-values < 0.05 statistically significant.

### Receiver-Operating Characteristic (ROC) Analysis

To anchor FST thresholds to functional vision, we performed ROC analyses. We defined binary outcomes as zero versus nonzero scores for each functional vision task: the Door Task and Line Task from the Mobility Test, and Table Test A① (tableware recognition), A② (figure recognition), B① (visual perception), and B② (visual exploration and relative position). The predictor variable was the FST threshold of the better-seeing eye (log cd·s/m²). The better-seeing eye was defined as the eye with better VA; if VA was identical in both eyes, the eye with the lower (better) FST threshold was selected. For each task, we calculated the area under the ROC curve with 95% confidence intervals. We determined the optimal FST cutoff value using the Youden index and reported sensitivity, specificity, and likelihood ratios.

## Results

### Participant Characteristics

We enrolled thirty-five patients (70 eyes; median age, 62 years; range, 39–84), comprising 16 males and 19 females. The distribution of VA categories was: NLP, 14 eyes; LP, 32 eyes; HM, 19 eyes; and CF or better (≥ 0.01), 5 eyes (**Table 1; Supplemental Table**). The median interval between visits was 5.6 weeks (range, 1.4–16.0 weeks). All enrolled patients completed all study examinations. We observed no testing-related adverse events.

**Table 1.** Demographics of subjects by visual acuity group.

| VA group | Eyes (n) | Patients (n)* | Male (n) | Female (n) | Median age (years) | Age range (years) |
| --- | --- | --- | --- | --- | --- | --- |
| NLP | 14 | 4 | 3 | 1 | 76.5 | 52.0–80.0 |
| LP | 32 | 15 | 6 | 9 | 63.0 | 49.0–84.0 |
| HM | 19 | 11 | 4 | 7 | 62.0 | 44.0–76.0 |
| ≥ CF | 5 | 5 | 3 | 2 | 50.0 | 39.0–82.0 |
| Total | 70 | 35 | 16 | 19 | 62.0 | 39.0–84.0 |

\*: the better eye's visual acuity was selected for counting the number when the two eyes' visual acuity groups differed within one subject.
VA: visual acuity, NLP: no light perception, LP: light perception, HM: hand motion, CF: counting fingers.

### FST Threshold Across VA Groups and Test-Retest Variability

Across all pooled visits, the median FST threshold was -0.451 log cd⋅s/m² (range, -6.241–2.909). **Supplemental Table** lists per-subject means across the three visits. The FST thresholds differed significantly across the VA categories (Kruskal–Wallis, *p* < 0.001; **Fig. 1**; **Table 2**). Median (range) thresholds by VA category were as follows: NLP, 1.13 (-0.63 to 2.54) log cd⋅s/m²; LP, -0.27 (-2.70 to 2.91) log cd⋅s/m²; HM, -1.13 (-6.24 to 0.51) log cd⋅s/m²; and CF or better: -2.82 (-5.67 to -1.73) log cd⋅s/m². Within-group variability was substantial, with thresholds spanning more than 3 log units among eyes in the same VA category. Notably, we obtained measurable FST responses in 9 of 14 NLP eyes (64.3%), highlighting FST’s ability to detect residual visual sensitivity even in patients without clinically detectable light perception. Post-hoc Dunn tests with Bonferroni adjustment showed significant pairwise differences for all comparisons except NLP vs. LP (adjusted *p* = 0.144) and HM vs. CF or better (*p* = 0.552); LP vs. HM (*p* = 0.018), LP vs. CF or better (*p* < 0.001), NLP vs HM (*p* < 0.001), and NLP vs. CF or better (*p* < 0.001) were significant.

**Figure 1.**
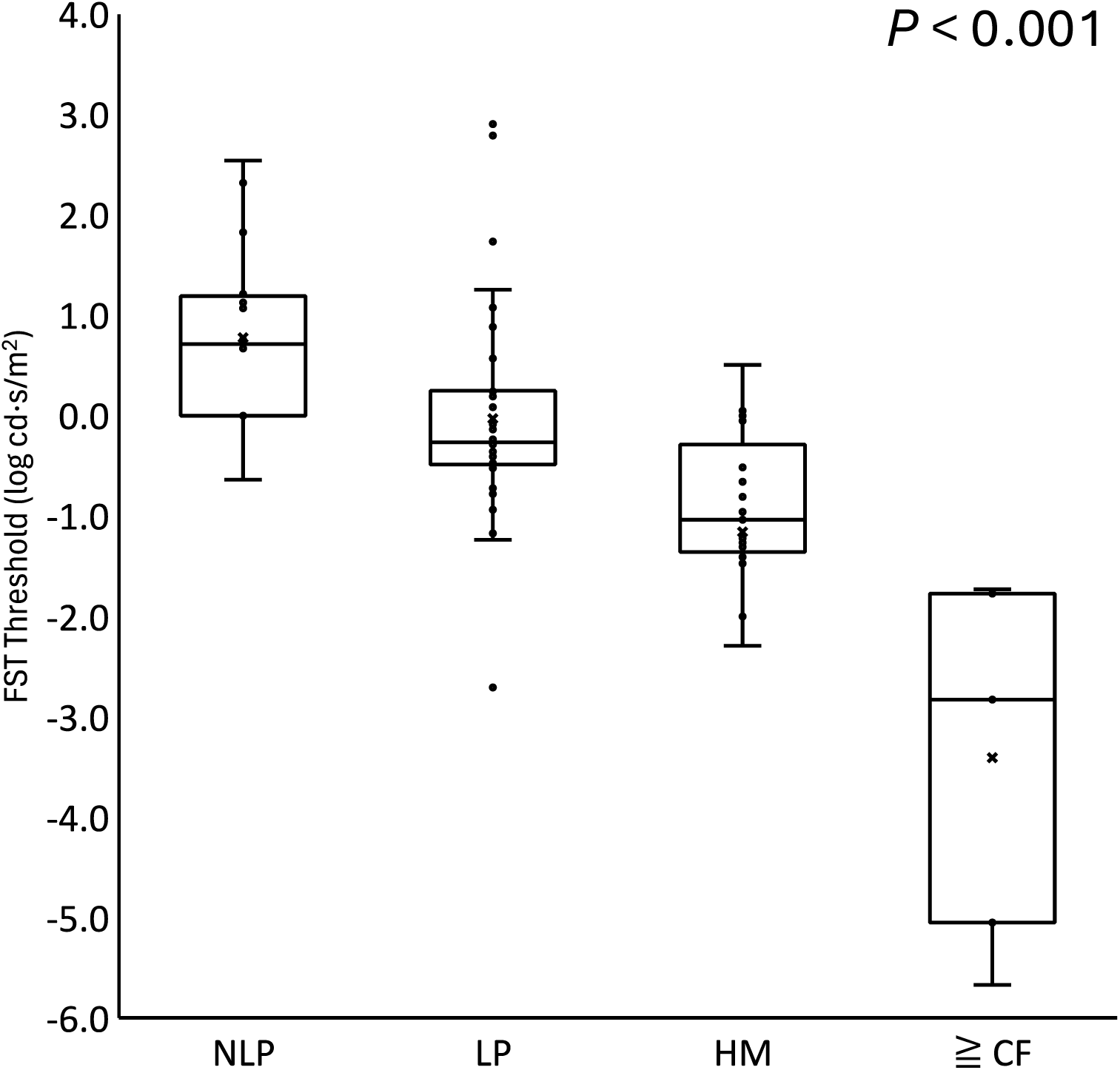
The Distribution of FST Thresholds in Each Visual Acuity Category. The ends of the boxes represent the upper and lower quartiles. The middle line in the box is the median value. Maximum and minimum values are shown as the upper and lower ends of the whiskers. FST thresholds differed significantly across VA categories (Kruskal–Wallis, p < 0.001). Post-hoc Dunn tests with Bonferroni adjustment showed significant pairwise differences for all comparisons except NLP vs LP (adjusted p = 0.144) and HM vs CF or better (p = 0.552); LP vs HM (p = 0.018), LP vs CF or better (p < 0.001), NLP vs HM (p < 0.001), and NLP vs CF or better (p < 0.001) were significant. NLP: no light perception, LP: light perception, HM: hand motion, CF: counting fingers, FST: full-field stimulus test.

**Table 2.** Results of Examinations by Visual Acuity (VA) Group.

| VA group | NLP | LP | HM | ≥ CF | P value |
| --- | --- | --- | --- | --- | --- |
| Number of eyes | 14 | 32 | 19 | 5 |  |
| FST threshold (log cd·s/m<sup>2</sup>) | 1.13*<br>(-0.63 ~ 2.54) | -0.27<br>(-2.70 ~ 2.91) | -1.13<br>(-6.24 ~ 0.51) | -2.82<br>(-5.67 ~ -1.73) | *P* < 0.001 |
| Table test score |  |  |  |  |  |
| Table test A-tableware<br>[% with score = 0] | 0<br>(0 ~ 0)<br>[4/4 (100%)] | 0<br>(0 ~ 4.83)<br>[10/15 (66.7%)] | 3.33<br>(0 ~ 7.17)<br>[2/11 (18.2%)] | 7.83<br>(6.67 ~ 8)<br>[0/5 (0%)] | *P* < 0.001 |
| Table test A-figure<br>[% with score = 0] | 0<br>(0 ~ 0)<br>[4/4 (100%)] | 0<br>(0 ~ 0.17)<br>[14/15 (93.3%)] | 0<br>(0 ~ 1.00)<br>[6/11 (54.5%)] | 1<br>(0.17 ~ 1)<br>[0/5 (0%)] | *P* < 0.001 |
| Table test B-visual<br>perception<br>[% with score = 0] | 0<br>(0 ~ 0)<br>[4/4 (100%)] | 0<br>(0 ~ 3.00)<br>[13/15 (86.7%)] | 1.17<br>(0 ~ 2.5)<br>[5/11 (45.5%)] | 2.83<br>(2.4 ~ 3)<br>[0/5 (0%)] | *P* < 0.001 |
| Table test B-visual<br>exploration and relative<br>position<br>[% with score = 0] | 0<br>(0 ~ 0)<br>[4/4 (100%)] | 0<br>(0 ~ 1.00)<br>[13/15 (86.7%)] | 0.2<br>(0 ~ 2)<br>[5/11 (45.5%)] | 3.17<br>(1.6 ~ 3.33)<br>[0/5 (0%)] | *P* < 0.001 |
| Mobility test score |  |  |  |  |  |
| Door task<br>[% with score = 0] | 0<br>(0 ~ 0)<br>[4/4 (100%)] | 0<br>(0 ~ 0)<br>[15/15 (100%)] | 0<br>(0 ~ 6)<br>[9/11 (81.8%)] | 4<br>(0 ~ 6)<br>[1/5 (20.0%)] | *P* < 0.001 |
| Line task<br>[% with score = 0] | 0<br>(0 ~ 0)<br>[4/4 (100%)] | 0<br>(0 ~ 0)<br>[15/15 (100%)] | 0<br>(0 ~ 6)<br>[8/11 (72.7%)] | 4<br>(2 ~ 6)<br>[0/5 (0%)] | *P* < 0.001 |
| Questionnaire score |  |  |  |  |  |
| VFQ25 | 28.3<br>(18.9 ~ 31.9) | 28<br>(17.4 ~ 40.7) | 33.2<br>(17.5 ~ 52.6) | 30.9<br>(23.2 ~ 36.9) | *P* = 0.269 |
| IVI | 1<br>(0.35 ~ 1.61) | 1.29<br>(0.61 ~ 1.57) | 1.25<br>(0.57 ~ 1.93) | 1.15<br>(0.68 ~ 1.57) | *P* = 0.853 |
| IVI-VLV | 1.235<br>(0.25 ~ 2.11) | 1.22<br>(0.46 ~ 2.32) | 1.43<br>(0.61 ~ 2.68) | 1.36<br>(0.93 ~ 1.86) | *P* = 0.901 |

\*: Median (range) is shown.
NLP: no light perception, LP: light perception, HM: hand motion, CF: counting fingers, FST: full-field stimulus test, VFQ-25: the 25-item National Eye Institute Visual Function Questionnaire, IVI: the Impact of Vision Impairment Questionnaire, IVI-VLV: the Impact of Vision Impairment-Very Low Vision.

The median difference in FST thresholds between visit 1 and visit 2 was 0.18 log cd·s/m² (range, 0.01–1.88); between visit 2 and 3, 0.26 log cd·s/m² (range, 0.01–1.10); and between visit 1 and 3, 0.31 log cd·s/m² (range, 0.01–2.55). These differences were not statistically significant (*p* = 0.368, 0.550, and 0.664, respectively; Wilcoxon signed-rank test). Across the three scheduled visits, the proportion of eyes with FST threshold differences ≤ 0.5 log cd·s/m² was 48 (84.2%) between visit 1 and 2, 42 (73.7%) between visit 2 and 3, and 39 (68.4%) between visit 1 and 3.

To further characterize reproducibility, we generated Bland–Altman plots across visits (see Supplemental Fig. S1), which showed no systematic bias and narrow 95% limits of agreement (visit 1–2: −0.62 to +0.69; visit 2–3: −0.85 to +0.79 log cd·s/m²). The coefficient of repeatability (CoR) was ±0.66 and ±0.82 log cd·s/m² for visit 1–2 and visit 2–3, respectively, consistent with recently published RP-specific data.^21^ Test–retest reliability was excellent, with an intraclass correlation coefficient (two-way random-effects model, absolute agreement; single measures) of 0.962 (95% CI, 0.942–0.977; p<0.001) across the three visits. No significant learning effect was observed across the three visits (Friedman test, p = 0.846), supporting the robustness of repeated FST assessments.

### Functional Vision Tests and Vision-Related Questionnaires

Table 2 summarizes the results of the Table Test, Mobility Test, and vision-related questionnaires by VA category. Given that we performed all tests binocularly and daily visual function is typically dominated by the better-seeing eye, we assigned VA categories based on each participant’s better eye. All patients with NLP scored 0 on all tasks of the Table Test and Mobility Test, representing a floor effect in the lowest VA stratum. In contrast, most patients with CF or better achieved nonzero scores on all functional tests. Performance in the LP and HM groups was intermediate and heterogeneous, with a substantial proportion remaining at floor on one or more tasks. We observed significant between-group differences for all tasks of the Table Test and Mobility Test (Kruskal–Wallis, all p < 0.001). In contrast, none of the three vision-related questionnaires showed significant differences across VA groups (see Supplemental Fig. S2).

### Correlations Between FST and Functional Vision Tests

We used the better-seeing eye’s FST threshold to perform correlation analyses between FST and functional vision tests and questionnaires. We found significant associations for all Table Test and Mobility Test tasks (Spearman’s rank correlation): Table Test A-tableware, *r* = -0.697, *p* < 0.001; Table Test A-figure, *r* = -0.457, *p* = 0.007; Table Test B-visual perception, *r* = -0.583, *p* < 0.001; Table Test B-visual exploration and relative position, *r* = -0.656, *p* < 0.001. Mobility Test-Door Task, *r* = -0.649, *p* < 0.001; Line Task, *r* = -0.653, *p* < 0.001. In contrast, FST thresholds did not correlate with scores on any of the three vision-related questionnaires (**Fig. 2**; **Supplemental Fig. S2**).

**Figure 2.**
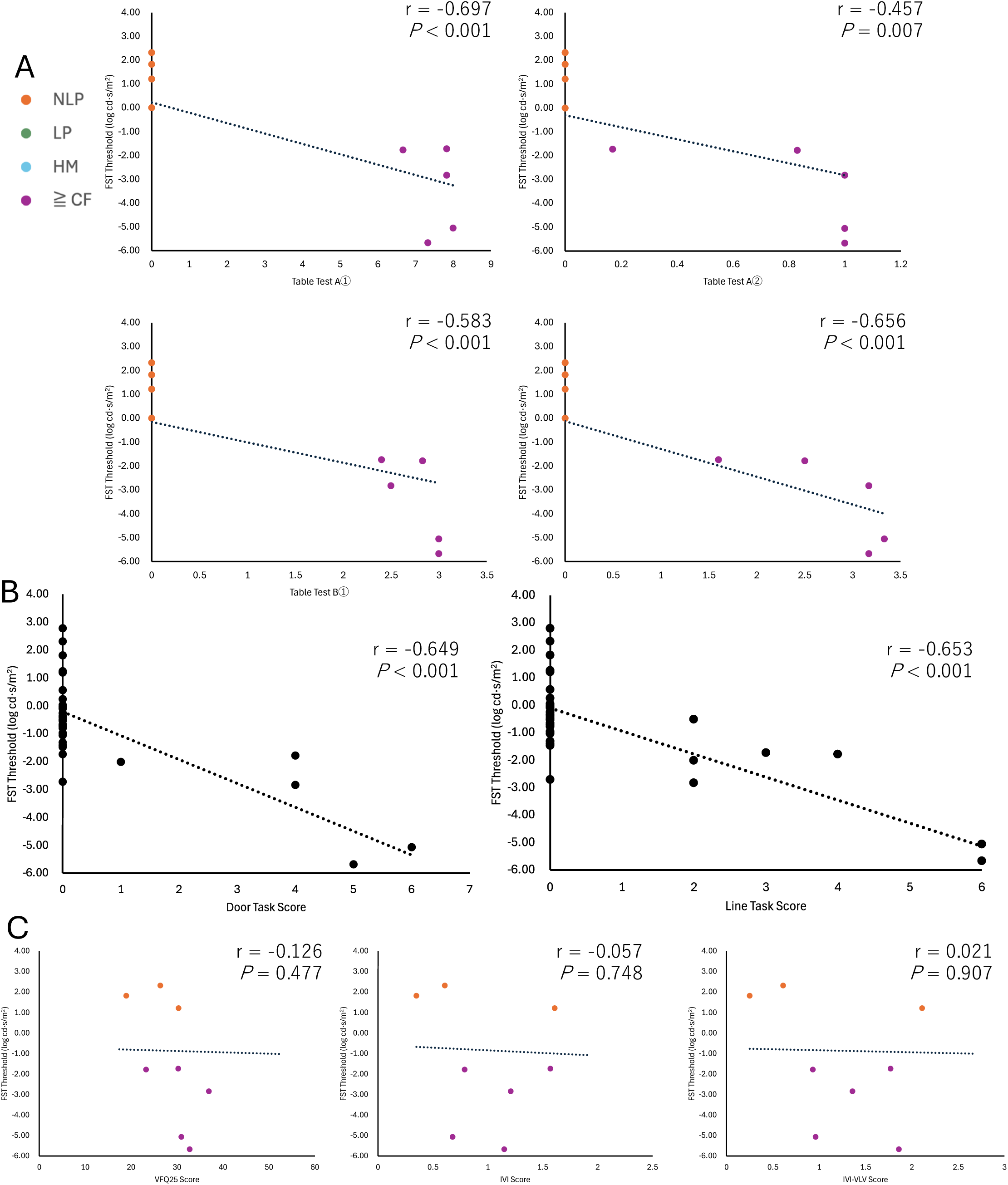
Correlations Between FST and Functional Vision Tests. **A. Correlation Between FST and Table Test.** Table Test A①: Table Test A-tableware, Table Test A②: Table Test A-figure, Table Test B①: Table Test B-visual perception, Table Test B②: Test B-visual exploration and relative position. **B. Correlation Between FST and Mobility Test.** **C. Correlation Between FST and Questionnaires.** All the association analyses were performed with Spearman’s rank correlation. NLP: no light perception, LP: light perception, HM: hand motion, CF: counting fingers, FST: full -field stimulus test.

### ROC Analysis for Transition From Zero to Nonzero Performance

To anchor FST thresholds at the point at which patients first demonstrated measurable functional vision, we conducted ROC analyses using a binary outcome of zero versus nonzero task performance (see Supplemental Fig. S3). For the Mobility Test, the AUROC was 0.99 (95% CI, 0.96–1.00; p<0.001) for the Door Task and 0.93 (95% CI, 0.83–1.00; p<0.001) for the Line Task. The Youden-optimal FST cutoffs were −1.75 log cd·s/m² for the Door Task (sensitivity 1.00, specificity 0.964) and −1.60 log cd·s/m² for the Line Task (sensitivity 0.88, specificity 0.962). For the Table Tests, discrimination ranged from acceptable to good. For Table Test A, the AUROC was 0.87 (95% CI, 0.76–0.99; p<0.001) for tableware recognition and 0.76 (95% CI, 0.57–0.95; p=0.009) for figure recognition. The corresponding Youden-optimal cutoffs were −0.87 log cd·s/m² (sensitivity 0.68, specificity 1.00) and −1.00 log cd·s/m² (sensitivity 0.73, specificity 0.83), respectively. For Table Test B, the AUROC was 0.78 (95% CI, 0.61–0.96; p=0.002) for visual perception and 0.85 (95% CI, 0.72–0.99; p<0.001) for visual exploration and relative position. Both Table Test B subtests yielded an identical Youden-optimal cutoff of − 0.87 log cd·s/m², with sensitivity 0.77 and specificity 0.86 for each. These cutoffs represent the FST levels at which patients who previously scored zero began to achieve successful task performance, highlighting the clinical relevance of FST improvements in enabling basic activities of daily living.

## Discussion

Current studies position FST as a promising psychometric tool for assessing efficacy in clinical trials for IRDs. In this ULV RP cohort, FST differentiated VA categories (NLP, LP, HM, CF, or better) and revealed substantial heterogeneity within categories, with eyes sharing the same off-chart VA label showing markedly different thresholds that could range over 3 log units. This within-group dispersion supports the use of FST as a more granular functional staging tool. Furthermore, we associated FST thresholds with functional vision in daily life, as reflected by the Table Test and Mobility Test, indicating that FST captures aspects of real-world visual capability beyond conventional acuity.

Categorical improvements, such as transitions from NLP to LP or from LP to HM, are intuitive and clinically meaningful; however, our findings highlight substantial variability within each VA category. This suggests that categorical endpoints and continuous FST thresholds can serve complementary roles: categorical changes provide patient- and clinician-friendly markers of vision gain, whereas FST offers a more granular, quantitative assessment of residual function and subtle changes within categories. These approaches strengthen the framework for evaluating efficacy in ULV clinical trials.

Among current treatments, vision restoration therapies—such as optogenetics, iPSC-based interventions, and retinal prostheses—primarily target patients with ULV, particularly those with NLP or LP. However, few studies have systematically evaluated FST in ULV cohorts, especially among NLP/LP. Jacobson et al.^12^ evaluated 7 NLP/LP participants; only one NLP participant was included, and FST was undetectable, with testing limited to red and blue stimuli. In our cohort, 14 NLP eyes were included; 9 (64.3%) yielded measurable FST thresholds spanning ∼3.2 log units, indicating that FST can quantify residual visual function even in NLP and may serve as a valid endpoint in ULV trials. This ability to obtain quantifiable thresholds in a majority of NLP eyes suggests that FST can expand and stratify the evaluable population in ULV trials, including patients who would otherwise be considered unmeasurable by conventional acuity tests.

In trials for RPE65-associated IRDs^7^, the Multi-Luminance Mobility Test has served as the primary outcome, approximating real-world navigation under standardized illuminance. While informative, it can be limited by logistical constraints and range effects (potential ceiling or floor effects at illuminance extremes)^22^. By contrast, FST is repeatable and feasible in standard clinic settings, with a wide dynamic range and minimal dependence on fixation. Furthermore, FST has been standardized under ISCEV and IPS guidelines, enhancing its comparability and facilitating implementation across multiple sites in global multicenter trials. Notably, in Japan, FST was adopted as a primary outcome measure in a Phase 3 trial of RPE65 gene therapy—the first such application worldwide^4^. In a prior study by Pierce et al.,^13^ FST correlated with the Ora-VNC navigation score. Taken together with our findings, these results suggest complementary roles: performance-based tests of functional vision in daily life provide ecological and face validity, whereas FST offers scalable, clinic-ready quantification across a broad sensitivity range.

Regulatory agencies emphasize that efficacy endpoints in vision restoration trials should reflect ADL and QoL^7,8,20^. In this study, we designed the activity-based tasks, namely the Mobility Test and the Table Test, to model ADL-relevant functions, including navigation and object recognition. The significant correlations between FST and these tasks demonstrate that FST can be anchored to outcomes directly linked to patient independence and daily functioning. These findings support the positioning of FST as a clinically meaningful endpoint within regulatory frameworks.

In this study, we did not find the significant difference in scores on patient-reported questionnaires (NEI VFQ-25, IVI, IVI-VLV) across the VA categories, nor did we identify their significant correlations with FST. Several factors likely contributed. First, adaptation and “response shift”—where patients recalibrate expectations and strategies over time-can attenuate associations between objective sensitivity and self-reported difficulty, especially in long-standing ULV. Both qualitative and quantitative studies on visual impairment document substantial psychosocial adjustment and coping mechanisms that buffer patient-reported impact despite profound vision loss^23,24^. Second, instrument targeting matters: the NEI VFQ-25 has known psychometric limitations in low-vision populations and was not designed for patients with very low vision; Rasch analyses show multidimensionality and misfit across several subscales in low-vision samples^25^. IVI-VLV was specifically developed and validated for severe vision loss and mitigates some of these issues, yet even targeted instruments may under-differentiate among ULV patients who have established stable routines and assistive strategies^20^. Notably, when visual function truly improves with therapy, PRO changes are often detectable. In neovascular AMD trials, the NEI VFQ-25 was responsive to VA gains, supporting its sensitivity to clinically meaningful improvement^26^. Therefore, PRO-FST links may become evident longitudinally in interventional settings even if cross-sectional baseline correlations are weak in ULV.

Visit-to-visit agreement in this cohort was favorable, with most eyes showing ≤ 0.5 log-unit differences. Recent RP-specific test–retest studies report minimal learning effects and repeatability coefficients of ±0.3–0.6 log units, providing an empirical yardstick for the smallest detectable individual change. Changes of ≳ 0.5–1.0 log units are plausible indicators of real improvement or decline at the eye level and align with thresholds used in IRD efficacy frameworks^21^.

Beyond reproducibility, establishing clinically meaningful change thresholds is critical for positioning FST as an efficacy endpoint. Our results, together with prior work, suggest that changes of ∼0.5 log cd·s/m² may represent the smallest detectable difference, while improvements of ≥1.0 log cd·s/m² can be interpreted as clinically meaningful. This threshold is supported on a distribution basis: the CoR of ±0.66 to ±0.82 log cd·s/m² establishes the upper boundary of measurement variability, and a ≥1.0 log-unit change exceeds this boundary, indicating a true signal beyond test–retest noise.

Furthermore, ROC analyses indicated that FST thresholds around –1.75 to –0.87 log cd·s/m² corresponded to the point at which patients first achieved nonzero performance in mobility and tabletop tasks, directly anchoring FST improvements to daily-life functionality. Such thresholds are consistent with responder definitions adopted in prior IRD gene therapy trials. Responder analyses stratified by baseline VA categories (NLP, LP, HM, CF) may provide a more nuanced interpretation. For example, a categorical shift from NLP to LP may be more functionally relevant than an equivalent continuous change. Anchoring these definitions to performance-based measures, such as mobility or tabletop tasks, would further strengthen the clinical meaningfulness of FST improvements.

Several limitations merit consideration. First, the CF or better subgroup was small (n = 5 eyes), which may limit statistical power and the precision of subgroup estimates. Second, we administered the Table Test and Mobility Test binocularly, whereas FST was measured monocularly; binocular summation and inter-ocular asymmetry could therefore introduce discordance across measures. Third, although repeated measurements were available, our analyses emphasized cross-sectional associations rather than within-person change. Future interventional studies should harmonize binocular or monocular conditions and anchor FST changes to performance-based functional vision and to PROs.

In conclusion, FST provides a granular, quantitative assessment of visual function in ULV, relates to functional vision in daily life, and is readily standardized and scalable. These findings strongly support FST as a candidate primary or supportive efficacy endpoint in clinical trials for RP patients with ULV.

## Data Availability

All data produced in the present study are available upon reasonable request to the authors

## Acknowledgments

We thank Naoko Nishimura, PhD, and Akiko Kamiya for their support of this clinical study, the orthoptists at Keio University Hospital for data collection, and Liz Messersmith, PhD, and David Hebert, PhD, for their advice on data analysis.

Declaration of Generative AI and AI-assisted technologies in the writing process: During the preparation of this work the authors used ChatGPT-4 in order to improve the readability and language of the manuscript. After using this tool, the authors reviewed and edited the content as needed and take full responsibility for the content of the publication.

## Meeting Presentation

Some data of this study was presented at the Association for Research in Vision and Ophthalmology (ARVO) annual meeting 2025 and has been submitted to ARVO 2026.

## Address for reprints

Dr. Toshihide Kurihara, Laboratory of Photobiology, Department of Ophthalmology, Keio University School of Medicine, 35 Shinanomachi, Shinjuku-ku, Tokyo 160-8582, Japan.

Dr. Yusaku Katada, Laboratory of Photobiology, Department of Ophthalmology, Keio University School of Medicine, 35 Shinanomachi, Shinjuku-ku, Tokyo 160-8582, Japan.

## List of Abbreviations

ADL: Activities of Daily Living
AMD: Age-Related Macular Degeneration
ANOVA: Analysis of Variance
CF: Counting Fingers
CoR: Coefficient of Repeatability
FST: Full-Field Stimulus Test
HM: Hand Motion
IPS: Imaging and Perimetry Society
IRD: Inherited Retinal Dystrophy
ISCEV: International Society for Clinical Electrophysiology of Vision
IVI: Impact of Vision Impairment Questionnaire
IVI-VLV: Impact of Vision Impairment–Very Low Vision Questionnaire
LogMAR: Logarithm of the Minimum Angle of Resolution
LP: Light Perception
NLP: No Light Perception
NEI VFQ-25: National Eye Institute Visual Function Questionnaire–25
QoL: Quality of Life
ROC: Receiver-Operating Characteristic
RP: Retinitis Pigmentosa
ULV: Ultra-Low Vision
VA: Visual Acuity

