## Supplemental Information for "Full-Field Stimulus Test for Visual Function Assessment in Ultra-Low Vision with Retinitis Pigmentosa"

### <u>The Protocol of Table Test</u>

#### 1. Table test A-tableware

Testing is conducted binocularly in a room at approximately 1000 lx (±30%). The table (85 × 85 cm) is covered with a black cloth (surface luminance 0.5 cd/m²), and white tableware items have luminance 140–150 cd/m². After a brief explanation and one training trial, the viewing distance is fixed at 60 cm. With eyes closed, 2–4 items from the predefined set are placed at predefined positions; the participant then opens the eyes and reports the total number, types, and locations (e.g., left/right/front/back). Response and completion time are recorded. Six trials are performed with varied placements. If items are not recognized, three trials are used to confirm non-recognition.

Materials and scoring: The tableware set comprises four white items: a large plate (diameter 22 cm), a small plate (diameter 13 cm), a cup (diameter 8.3 cm), and a spoon (length 16 cm). Each trial is scored on three components: (1) number of items—2 points if the participant correctly reports the total count, 0 points otherwise; (2) location—1 point for each item whose position on the table is correctly identified; and (3) type—1 point for each correctly identified item type, regardless of whether its position is also correct. The per-trial score is the sum of these components, and the final Table Test A-tableware score is the mean of the per-trial scores across all six trials.

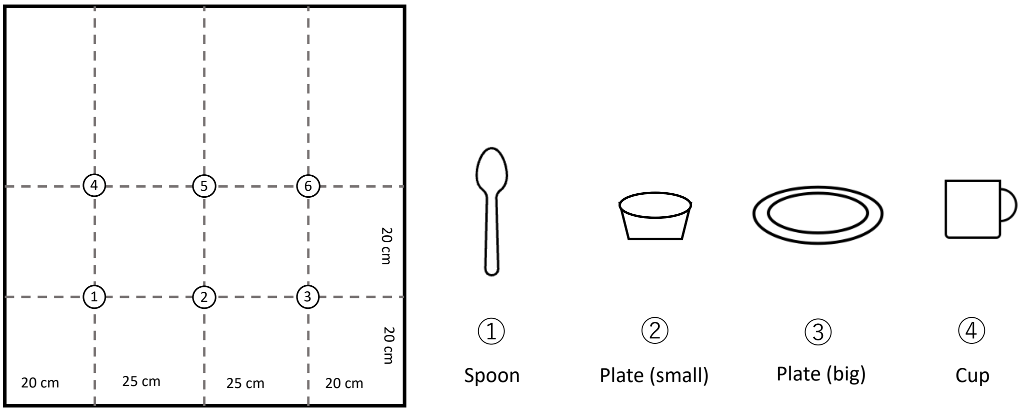


#### 2. Table test A-figure

Under the same lighting and setup (room ~1000 lx (±30%); black tablecloth 0.5 cd/m²; viewing distance 60 cm), a single high-contrast geometric shape (triangle, circle, square or rectangle) is presented on the tabletop. After instructions, the participant names the shape; correctness and completion time are recorded. Six randomized trials are completed; if shapes are not recognized, three confirmatory trials are performed.

Materials and scoring: Four white geometric shapes of approximately 8 × 8 cm are used: circle (diameter 4 cm), square (side 4.5 cm), triangle (4.5 × 4.5 cm), and rectangle (2.4 × 6.5 cm). Each trial scores 1 if the participant correctly names the shape and 0 otherwise. The final Table Test A-figure score is the proportion of correct identifications across all six trials (range, 0–1).

#### 3. Table Test B – visual perception.

Testing is performed binocularly on a white tablecloth (~150 cd/m²) in a room at ~1000 lx (±30%), using objects of predefined contrast (black ≤ 1 cd/m²; gray ~ 50 cd/m²). With eyes closed, a single object is placed at a predefined left/center/right position; the participant then opens the eyes, reports presence and position, and touches the object (first touch adjudicates). Responses and completion time are recorded across six randomized trials; if the object is not perceived, three trials confirm non-recognition.

Materials and scoring: Objects of two contrast levels are used—black (luminance ≤ 1 cd/m²) and gray (luminance approximately 50 cd/m²)—including notebooks (B6 size) and staple boxes. Each trial is scored on three components: (1) recognition—1 point if the participant reports the object's presence; (2) position—1 point if the location (left, center, or right) is correctly identified; and (3) touch—1 point if the participant's first touch contacts the object. The maximum score per trial is 3, and the final Table Test B-visual perception score is the mean across all six trials (range, 0–3).

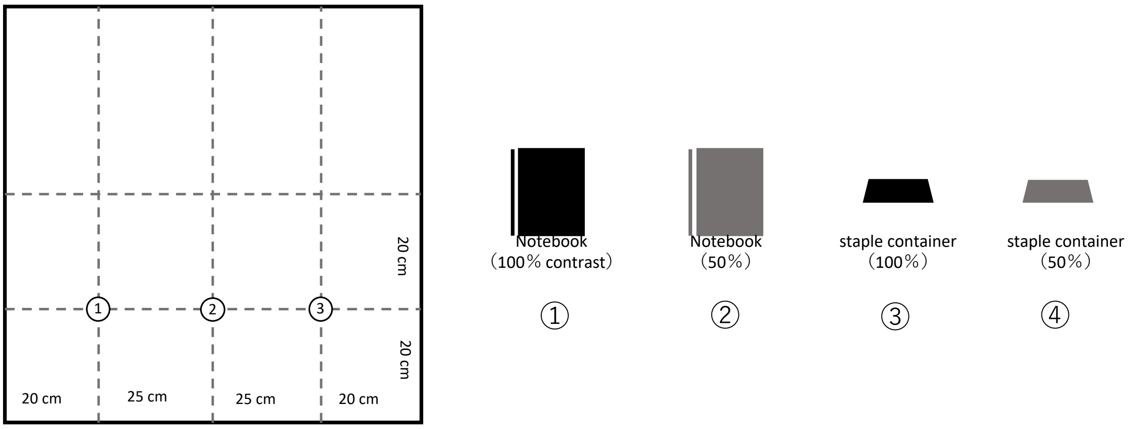


#### 4. Table Test B – visual exploration and relative position.

With the same environment and viewing distance (white tablecloth ~150 cd/m²; room ~1000 lx (±30%); 60 cm), two or three identical-contrast tumblers are placed at predefined locations. After opening the eyes, the participant reports the number of tumblers and their table positions (verbal report or pointing permitted). Responses and completion time are recorded. Six trials are performed with varied placements; non-recognition is verified by three trials.

Materials and scoring: Tumblers of uniform contrast (black or gray) are used, with two or three placed per trial. Each trial is scored on two components: (1) number—1 point if the total count is correct, 0 otherwise; and (2) position—1 point for each tumbler whose location is correctly identified. The maximum per-trial score is 3 (two tumblers: 1 + 2) or 4 (three tumblers: 1 + 3). The final Table Test B-visual exploration and relative position score is the mean across all six trials.

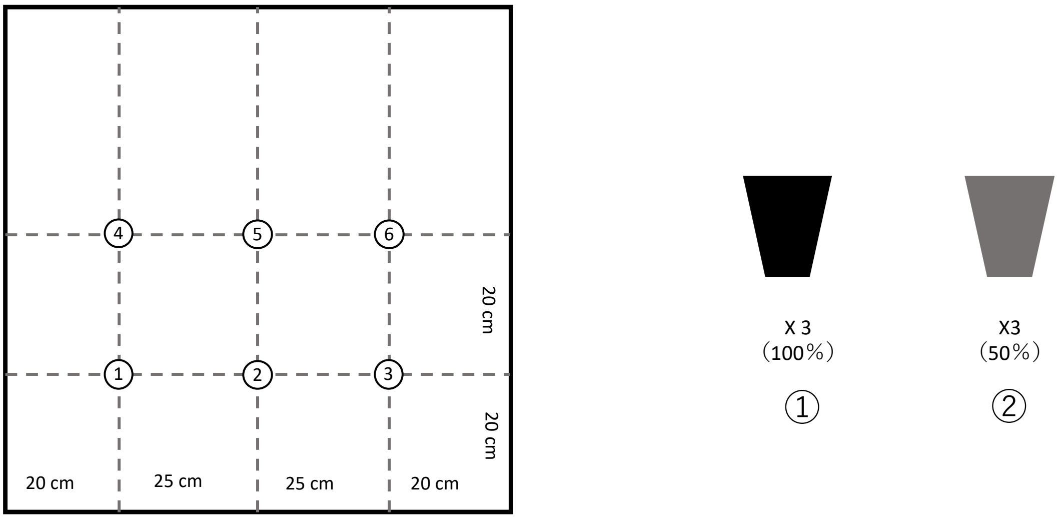

<u>**The Protocol of Mobility Test**</u>

Environment (applies to both tasks).

Testing is performed in a 3.35 × 5.35 m room with white walls (white cloth) and a black floor. Room illuminance is controlled and tested in sequence at approximately 400, 250, 100, 50, 10, and 1 lux (tolerance: 250 and 400 lx, ±30%; 50 and 100 lx, ±50%; 1 and 10 lx, ±200%), beginning at 400 lux and progressing to lower levels while criteria are met. Each illuminance level consists of six trials; start positions (three) and target positions are randomized per a predefined plan. All sessions are video-recorded; completion time is recorded for every trial.

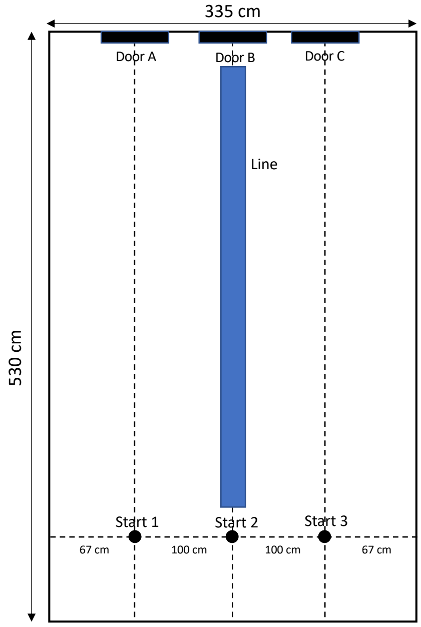


**Mobility Test – Door Task (with scoring).**

A rectangular "door" (55 × 100 cm; upper edge 200 cm above the floor) is displayed on the white wall. At each illuminance level, six randomized trials are completed. A trial is recorded as success if the participant walks from the start position to the door and touches any point within the door area; otherwise, failure is recorded along with the shortest perpendicular distance from the fingertip at touch to the nearest door edge. Testing proceeds from 400 lux to the next lower illuminance only if all six trials at the current level are recorded as success. The lowest luminance level which all 6 trials was succeeded was recorded as the result as the Door Task, and was converted to an ordinal score as follows: 0 points if 400 lux is not passed; 1 point if 400 lux is passed; 2 points if 250 lux is passed; 3 points if 100 lux is passed; 4 points if 50 lux is passed; 5 points if 10 lux is passed; and 6 points if 1 lux is passed. (Example: passing 50 lux but not 10 lux yields 4 points.) Report the luminance score, mean completion time, and any failure distances.

White-cane use is permitted and recorded. The examiner does not provide directional guidance to the participant. If the examiner judges that the participant is at risk of colliding with a wall, or if any part of the participant's body or clothing contacts a wall, the examiner verbally intervenes and suspends the trial; the trial is then resumed from the corrected position, and this intervention alone does not constitute a failure.

**Mobility Test – Line Task (with scoring).**

A single white line (10 × 439 cm) is placed along the center of the black floor. At each illuminance level, six randomized trials are completed using the predefined start positions. A trial is a success if the participant stops within 30 cm of the line's distal end with at least part of one foot resting on the line; otherwise, failure is recorded together with the shortest distance from the participant's feet to the line end. The Line Task uses the same luminance progression and scoring rules as the Door Task. At each illuminance level, participants complete six trials; a level is considered passed only if all six are successful, and the luminance score is assigned using the same scheme as the Door Task.

White-cane use is not permitted during the Line Task. The examiner does not provide directional guidance. If the examiner judges that the participant is at risk of colliding with a wall, or if any part of the participant's body or clothing contacts a wall, the examiner verbally intervenes and suspends the trial; the trial is then resumed from the corrected position, and this intervention alone does not constitute a failure.
