## Supplemental Figure S1 for "Full-Field Stimulus Test for Visual Function Assessment in Ultra-Low Vision with Retinitis Pigmentosa"

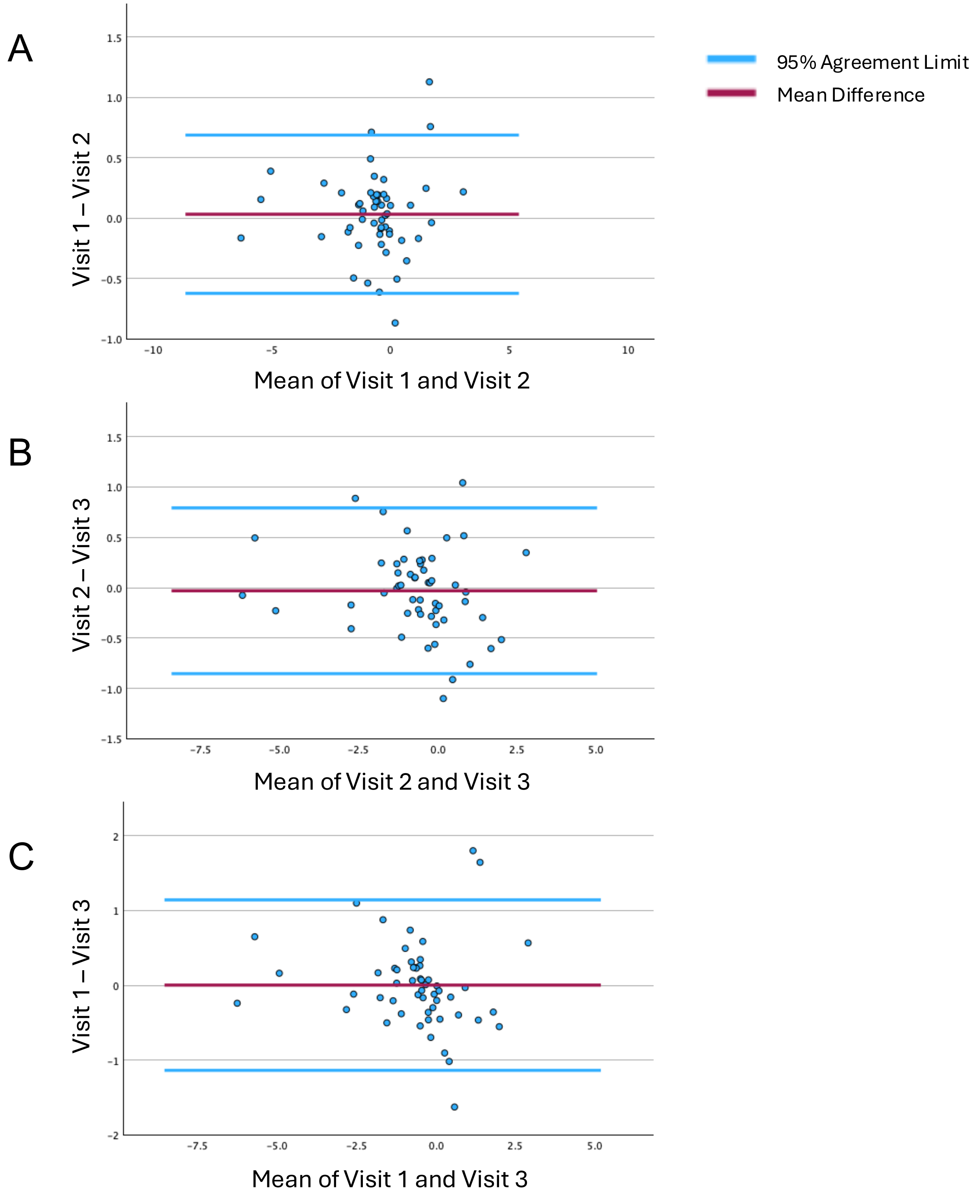


**Supplemental Figure S1. Bland–Altman plots of FST reproducibility across visits.**
(A) Visit 2 versus Visit 1, (B) Visit 3 versus Visit 2, and (C) Visit 3 versus Visit 1. Differences are plotted against the mean of the two visits. Solid horizontal lines indicate the mean bias, and dashed lines indicate the 95% limits of agreement (bias ± 1.96 SD). Mean bias and 95% limits of agreement were: (A) 0.03 (−0.62 to 0.69), (B) −0.03 (−0.86 to 0.80), and (C) 0.002 (−1.14 to 1.14) log cd·s/$m^2$. The corresponding coefficients of repeatability (CoR) were (A) ±0.66, (B) ±0.82, and (C) ±1.14 log cd·s/$m^2$.
