## Supplemental Figure S2 for "Full-Field Stimulus Test for Visual Function Assessment in Ultra-Low Vision with Retinitis Pigmentosa"

A

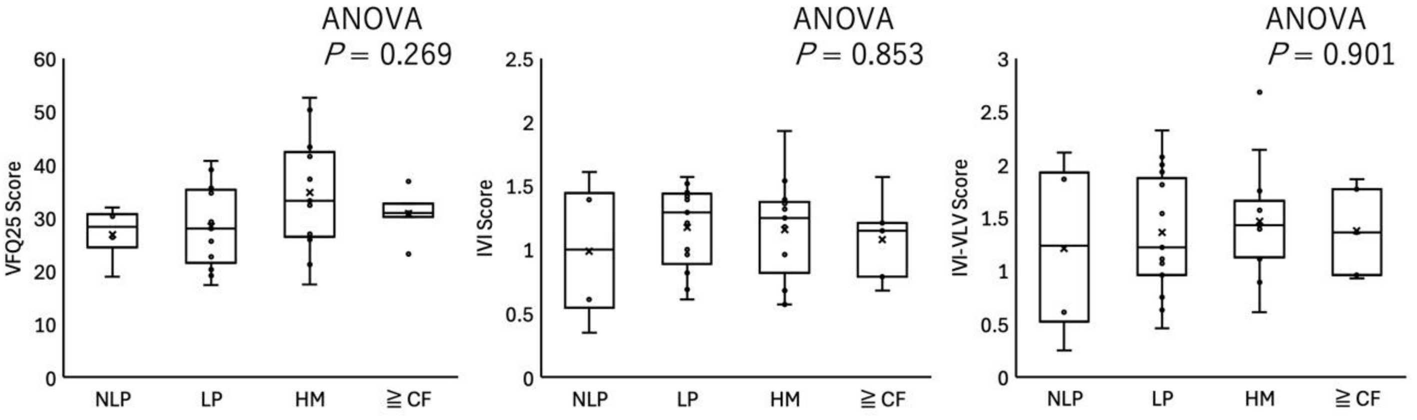


B

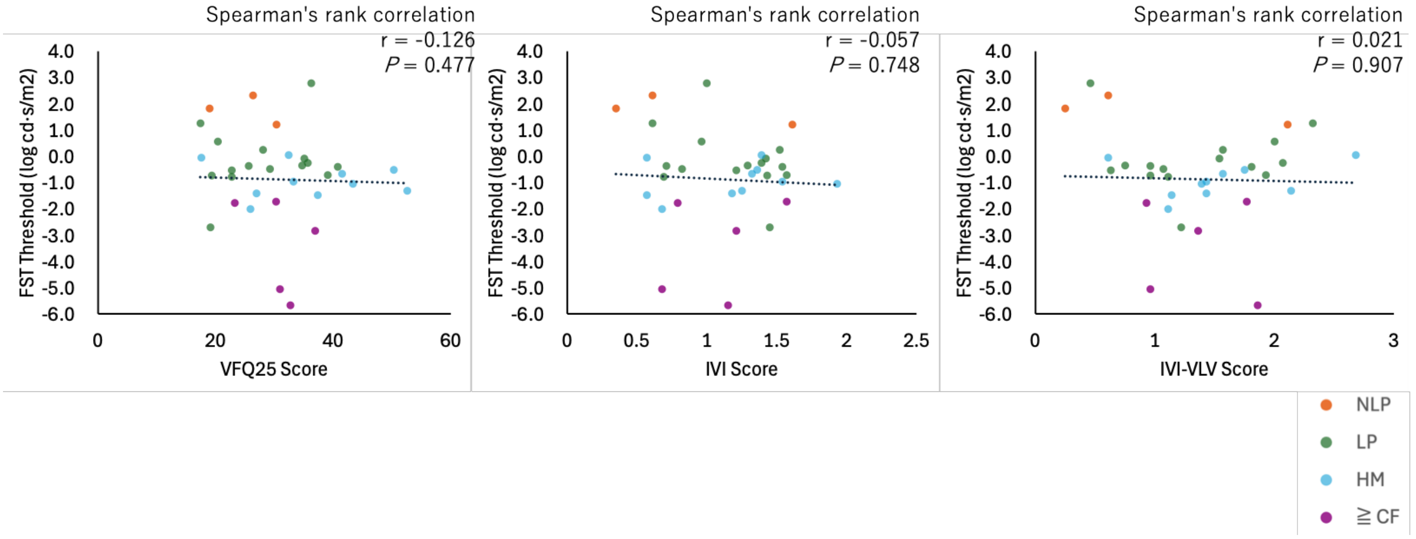


**Supplemental Figure S2**. (A) Vision-related quality-of-life questionnaire scores by VA category. Boxes indicate the interquartile range (IQR), the center line denotes the median, and whiskers span the minimum and maximum values. No significant differences were detected across VA categories. (B) Relationship between FST threshold (log cd/m$^2$) and questionnaire scores (NEI VFQ-25, IVI, IVI-VLV). No significant correlations were identified.
CF, counting fingers; HM, hand motion; LP, light perception; NLP, no light perception; FST, full-field stimulus test; VA, visual acuity; NEI VFQ-25, 25-item National Eye Institute Visual Function Questionnaire; IVI, Impact of Vision Impairment; IVI-VLV, Impact of Vision Impairment–Very Low Vision.
