## Supplemental Figure S3 for "Full-Field Stimulus Test for Visual Function Assessment in Ultra-Low Vision with Retinitis Pigmentosa"

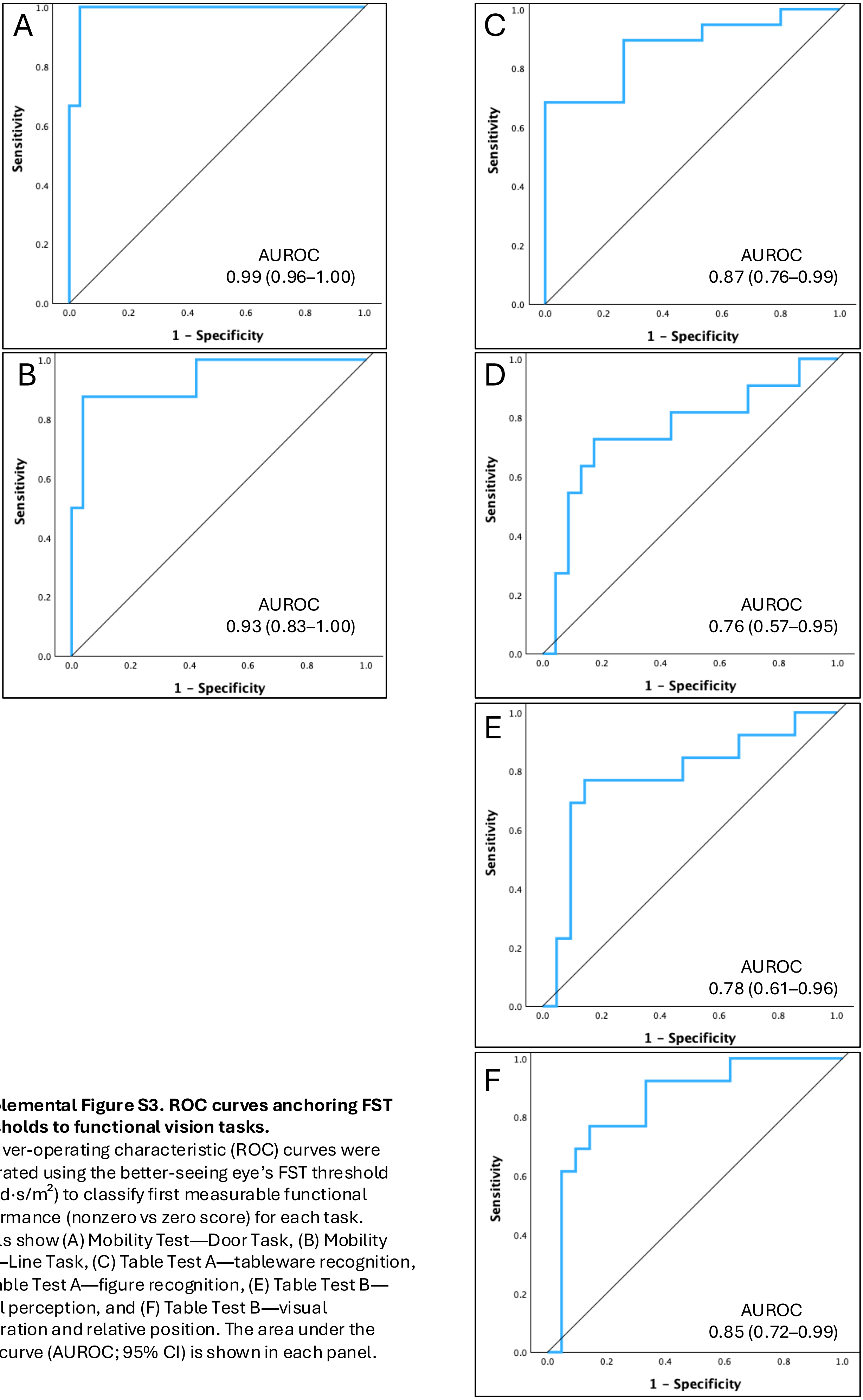


**Supplemental Figure S3. ROC curves anchoring FST thresholds to functional vision tasks.**
Receiver-operating characteristic (ROC) curves were generated using the better-seeing eye's FST threshold (log cd·s/$m^2$) to classify first measurable functional performance (nonzero vs zero score) for each task. Panels show (A) Mobility Test—Door Task, (B) Mobility Test—Line Task, (C) Table Test A—tableware recognition, (D) Table Test A—figure recognition, (E) Table Test B—visual perception, and (F) Table Test B—visual exploration and relative position. The area under the ROC curve (AUROC; 95% CI) is shown in each panel.
