## Supplemental Table for "Full-Field Stimulus Test for Visual Function Assessment in Ultra-Low Vision with Retinitis Pigmentosa"

Supplemental table. FST Thresholds of Each Subject

| Patient No. | Sex | Age (range; years) | VA | | VA Category | | FST Threshold (log cd·s/m$^2$)* | |
|---|---|---|---|---|---|---|---|---|
| | | | RE | LE | RE | LE | RE | LE |
| P001 | Male | 46-50 | LP | LP | LP | LP | 0.259 | 0.245 |
| P002 | Male | 41-45 | 0.03 | HM | ≧ CF | HM | -1.773 | -1.219 |
| P003 | Female | 41-45 | HM | HM | HM | HM | -1.262 | -1.469 |
| P004 | Female | 81-85 | HM | 0.15 | HM | ≧ CF | -2.293 | -5.667 |
| P005 | Female | 76-80 | HM | HM | HM | HM | N/A | -2.000 |
| P006 | Female | 61-65 | LP | LP | LP | LP | -0.133 | -0.250 |
| P007 | Female | 66-70 | HM | NLP | HM | NLP | -1.036 | 1.129 |
| P009 | Female | 66-70 | LP | LP | LP | LP | -2.704 | -1.232 |
| P010 | Male | 71-75 | NLP | LP | NLP | LP | 0.669 | -0.705 |
| P012 | Female | 56-60 | HM | LP | HM | LP | -1.403 | -1.171 |
| P013 | Female | 46-50 | LP | LP | LP | LP | -0.395 | 0.196 |
| P014 | Male | 56-60 | LP | LP | LP | LP | -0.461 | -0.776 |
| P015 | Female | 51-55 | LP | LP | LP | LP | -0.470 | -0.438 |
| P016 | Male | 41-45 | HM | LP | HM | LP | -0.517 | -0.236 |
| P017 | Female | 51-55 | NLP | NLP | NLP | NLP | N/A | 2.322 |
| P018 | Male | 51-55 | NLP | LP | NLP | LP | 1.074 | -0.354 |
| P019 | Male | 76-80 | NLP | NLP | NLP | NLP | 1.827 | -0.635 |
| P020 | Female | 61-65 | NLP | LP | NLP | LP | 2.539 | -0.086 |
| P021 | Female | 61-65 | LP | HM | LP | HM | -0.410 | -0.658 |
| P022 | Male | 66-70 | 0.03 | HM | ≧ CF | HM | -1.725 | -1.284 |
| P023 | Female | 61-65 | LP | LP | LP | LP | -0.521 | -0.282 |
| P024 | Male | 66-70 | LP | HM | LP | HM | 0.084 | -0.047 |
| P025 | Male | 51-55 | LP | HM | LP | HM | 0.889 | 0.047 |
| P026 | Female | 61-65 | LP | LP | LP | LP | 2.909 | 2.794 |
| P027 | Female | 61-65 | HM | HM | HM | HM | 0.506 | -1.306 |
| P028 | Female | 36-40 | HM | CF | HM | ≧ CF | 0.004 | -2.825 |
| P029 | Female | 66-70 | LP | LP | LP | LP | 1.738 | 1.257 |
| P030 | Female | 81-85 | LP | LP | LP | LP | -0.340 | -0.204 |
| P031 | Female | 61-65 | HM | HM | HM | HM | -0.805 | -0.956 |
| P032 | Male | 76-80 | NLP | NLP | NLP | NLP | N/A | N/A |
| P034 | Male | 71-75 | NLP | NLP | NLP | NLP | 1.214 | N/A |
| P035 | Male | 71-75 | LP | LP | LP | LP | 1.077 | 0.574 |
| P036 | Male | 46-50 | LP | CF | LP | ≧ CF | -0.932 | -5.049 |
| P037 | Male | 56-60 | NLP | LP | NLP | LP | 0.755 | -0.719 |
| P038 | Male | 41-45 | NLP | HM | NLP | HM | N/A | -6.241 |

* The averaged value of the FST thresholds of three visits for each patient.

VA: visual acuity, RE: right eye, LE: left eye, NLP: no light perception, LP: light perception, HM: hand motion, CF: counting fingers, FST: full-field stimulus test.
